# Protecting women from economic shocks to prevent HIV in Africa: Evidence from the POWER randomised controlled trial in Cameroon

**DOI:** 10.1101/2024.02.02.24302170

**Authors:** Aurélia Lépine, Sandie Szawlowski, Emile Nitcheu, Henry Cust, Eric Defo Tamgno, Julienne Noo, Fanny Procureur, Illiasou Mfochive, Serge Billong, Ubald Tamoufe

## Abstract

**Background:** Women in Sub-Saharan Africa are disproportionately affected by the HIV epidemic. Young women are twice as likely to be living with HIV than men of the same age and account for 64% of new HIV infections among young people. Many studies suggest that financial needs, alongside biological susceptibility, are the main causes of the gender disparity in HIV acquisition. While the literature shows a limited understanding of the link between poverty and HIV, there is some new robust evidence demonstrating that women adopt risky sexual behaviours as a way to cope with economic shocks.

**Methods:** We recruited 1,506 adolescent girls and women engaging in transactional or in commercial sex in Yaounde, Cameroon, using snowball sampling. Half of the study participants were randomly allocated to receive a free health insurance product covering themselves and their economic dependents over 12 months. We collected data on socio-economics, health and sexual behaviours and sexually transmitted infection and HIV biomarkers at baseline, 6-month post randomisation (midline) and 12-month post randomisation (endline).

**Results:** We found that study participants engaging in transactional sex allocated to the treatment group were less likely to be infected with HIV (OR=0.109, p-value<0.05). We showed that the intervention allowed women to leave transactional sex. In addition, we found that for the participants remaining in transactional sex, the intervention increased condom use and reduced sex acts, these results were however only statistically significant at 10% given our sample size. There was no evidence of a change in risky sexual behaviours or in a reduction of HIV incidence among female sex workers.

**Conclusion:** The study provides the first evidence of the effectiveness of a formal shock-coping strategy to prevent HIV among women in Africa. We showed that the intervention effectiveness operates through the reduction in health shocks since the increase in healthcare use following the intervention was low. We estimated that in our trial, the cost for each HIV infection averted is £4,667 among the cohort of women engaging in transactional sex. Policymakers should consider formal shock-coping strategies to prevent HIV among women in Africa.

## 1 Introduction

Women aged 15-24 in Sub-Saharan Africa are disproportionally affected by the HIV epidemic: they are twice as likely to live with HIV than men of the same age and account for 63% of new HIV infections among young people (1). Today, new HIV infections are three times higher in young women than their male counterparts (2). In Cameroon, a country with one of the highest gender disparities in HIV globally, adolescent girls aged 15–19 are five times more likely to be infected with HIV than their male counterparts (2% versus 0.4%) (3). Globally, female sex workers are a key population in the fight against HIV and are now 26 times more likely to be infected than the general population, up from 13 times in 2017 (1,4). There is growing evidence that heterosexual sex as part of commercial and transactional relationships, *“non-commercial sexual relationships in exchange for material support and benefits”,* are a key driver of the HIV epidemic in sub-Saharan Africa (5,6).

Many studies suggest that financial needs, alongside biological susceptibility, are the leading causes of the gender disparity in HIV acquisition through the adoption of risky sexual behaviours by young women associated with commercial and transactional sex (7–9). Whilst the lives of women and girls have dramatically improved over the last quarter century, there has been limited progress towards gender equality for the world’s poorest, particularly amongst marginalised or women living in remote areas (10). Structural gender disparities mean women lack the same access as men to formal, well-paid jobs, education and productive assets (e.g. land), limiting their economic security. This reduces the ability of women, particularly non-married women, to deal with economic insecurity since they face barriers to accessing formalised coping strategies. Commercial and transactional sex is an attractive risk-coping strategy with the ability to raise quick money and earn up to three times alternative occupations (11). Not only that, the network of male partners can result in other financial or in-kind gifts used to cope with economic shocks (12). Within such relationships, women are incentivised to provide unprotected sex to men who desire it (13) because they can earn between 9% more in Kenya and 66% more in India for unprotected sex (14–16). Recent evidence suggests that income variability is more important than income level, with sudden economic shocks consistently leading to significant increases in risky sexual behaviours to provide their basic needs (17,18).

Previous structural interventions to address the link between poverty and HIV have been cash transfers. These are effective in raising income and raising the opportunity cost of engaging in risky transactional or commercial relationships, particularly when conditions for merit good are included, e.g. school attendance or remaining HIV negative. Evidence for effectiveness, however, is mixed. Only three of eight studies found them protective of HIV and STIs and only when conditions were set (19). It is hypothesised that low monetary values and poor targeting toward economic shocks are why such programmes are not building economic resilience as expected (17). Indeed, the scale appears important in achieving protection, with evidence of country-level cash programmes finding modest protection against HIV (20).

Building on this evidence, we implemented the POWER study (*Protecting women from economic shocks to fight HIV in Africa*) to fill this gap in knowledge by protecting women engaging in transactional and commercial sex against one the most common economic shocks experienced by households in sub-saharan Africa: health shocks (21). POWER is a randomised controlled trial that aims to reduce the impact of economic shocks by providing health insurance without co-payment to eliminate health expenses at the point of use for vulnerable women most at risk of HIV in Cameroon. Out of pocket medical spendings are highly costly, concentrated among the poor, and are the most prevalent in urban areas compared to other economic shocks (22). In urban Cameroon, 21% of households spend more than 10% of their income on health out-of-pocket spending every year, and only 2.7% of women and 8.2% of men have health insurance coverage (23). In this paper, we evaluate the effectiveness of this intervention in reducing economic shocks, risky sexual behaviours, HIV incidence and other STIs among women engaged in commercial or transactional sex in Yaundé, Cameroon.

## 2 Methods

A detailed description of the methods can be found in the accompanying protocol paper (24).

### 2.1 Recruitment and ethics

We conducted a stratified randomised controlled trial in Yaoundé Cameroon, a country chosen because of the highest HIV gender disparities in the world. The sample is stratified 1:1 by the type of risky sex activity: commercial or transactional sex. We identified two community-based organisations (CBO) working with these groups. The CBO “Renata” was chosen to recruit and follow women engaging in transactional sex, while “Horizon Femmes” was chosen for its expertise in working with female sex workers (FSWs). These sites were used for data collection, and we worked with CBO staff who interacted with the research team and the study participants. Participants in both commercial and transactional sex groups were recruited using snowball sampling, given that those populations were hidden, especially FSWs, due to sex work being illegal in Cameroon (25). The CBO initially identified 34 and 23 women engaging in transactional and commercial sex, respectively, who were seeds and had a large network in their community. Each of these seeds and subsequent recruited participants were asked to recruit up to three other women meeting the study eligibility criteria and received an incentive of 500 FCFA (£0.64) for each recruited participant. This led to, on average, 21 transactional and 31 commercial women being recruited per seed and to a total sample of 1,506 participants recruited in the study.

To be eligible to participate in the study, the participant needed to engage in transactional or commercial sex in Yaoundé, Cameroon, to be female, to be aged 15 or over, to not be married, to have at least one economic dependent living in Yaoundé (no family relationship required), to be HIV negative, to have a password protected phone and the ability to respond to SMS messages. STIs were conducted at baseline with free treatment offered to those testing positive and referral to the CBO if participants were severely depressed, victims of violence or victim of a recent rape.

Ethics approval was provided by UCL ethics committee (ref: 17341/00) and the National Ethics Committee in Cameroon (CNERSH, ref: 2020/12/1313).

### 2.2 Randomisation and blinding

Participant randomisation occurred between October and December 2021, i.e. two months after the baseline survey had taken place, and only those that returned for randomisation are included in the analysis. Participatory randomisation was found to be preferable to computer-based randomisation during the formative research phase as it was perceived to be more transparent and fair to respondents (26). Participants were told that they would all receive the intervention but that the treatment group would receive it the day of the randomisation, while the control group would have to wait 12 months to receive it. Randomisation was conducted in a private room and occurred in the presence of two enumerators and one survey supervisor. A description of the randomisation procedure checks for eligibility, and informed consent for randomisation took place. Parental consent was sought for minors under 21 years of age. Participants were then presented with a large, deep black bag and watched the enumerator place two coloured balls inside: one orange ball (for the treatment group) and one white ball (control group). To determine their group allocation, the participant draws a ball from the bag. Those picking the orange ball (treatment group) received a detailed description of the insurance product they would receive and were asked to re-consent to participate in the study. Those allocated to the control group were re-informed of all services they could still benefit from participation if they decided to remain in the study and were told again that they would receive the intervention at the end of the study if they continued to participate. In addition, they were offered counselling from the CBO to minimise frustration for not receiving the intervention immediately. Whilst counselling appeared popular in advance during the formative research, it was not widely taken up, with only 12% opting for it after allocation to the control group (26). Figure A0 shows the project timeline with dates for baseline, randomisation and the two follow-up surveys.

Table 1 demonstrates successful randomisation. There is a significant difference between respondents who have children in the commercial sex strata, but this is within the expectations of randomisation given the number of characteristics compared.

**Table 1:** Summary statistics at baseline.

|  | Commercial Sex |  |  |  |  | Transactional Sex |  |  |  |  |
| --- | --- | --- | --- | --- | --- | --- | --- | --- | --- | --- |
|  | Treatment |  | Control |  | p-val of the difference | Treatment |  | Control |  | p-val of the difference |
|  | Obs | Mean / % | Obs | Mean / % |  | Obs | Mean / % | Obs | Mean / % |  |
| Age, years | 286 | 31.2 | 287 | 31.0 | 0.790 | 290 | 25.1 | 278 | 24.3 | 0.114 |
| Experience - months in sex work | 289 | 56.6 | 290 | 54.9 | 0.691 | 289 | 38.8 | 278 | 36.0 | 0.395 |
| **Nationality** |  |  |  |  |  |  |  |  |  |  |
| Cameroonian (%) | 289 | 100% | 290 | 99% | 0.566 | 290 | 100% | 278 | 100% | 0.308 |
| **Education** |  |  |  |  |  |  |  |  |  |  |
| Currently studying | 289 | 14% | 290 | 19% | 0.099 | 290 | 44% | 278 | 49% | 0.290 |
| **Highest achieved level of education** |  |  |  |  |  |  |  |  |  |  |
| Primary | 289 | 15% | 290 | 14% | 0.801 | 290 | 8% | 278 | 7% | 0.517 |
| Secondary primary cycle | 289 | 36% | 290 | 30% | 0.105 | 290 | 24% | 278 | 25% | 0.774 |
| Secondary second cycle | 289 | 28% | 290 | 31% | 0.430 | 290 | 39% | 278 | 33% | 0.144 |
| Superior | 289 | 19% | 290 | 22% | 0.422 | 290 | 28% | 278 | 33% | 0.129 |
| No education | 289 | 2% | 290 | 3% | 0.404 | 290 | 0% | 278 | 1% | 0.296 |
| Other | 289 | 0% | 290 | 0% | 0.319 | 290 | 1% | 278 | 1% | 0.688 |
| **Marital status** |  |  |  |  |  |  |  |  |  |  |
| Never married | 289 | 89% | 290 | 89% | 0.803 | 290 | 97% | 278 | 98% | 0.645 |
| Separated | 289 | 7% | 290 | 7% | 0.860 | 290 | 2% | 278 | 2% | 0.946 |
| Divorced | 289 | 1% | 290 | 2% | 0.205 | 290 | 0% | 278 | 0% |  |
| Widowed | 289 | 3% | 290 | 2% | 0.789 | 290 | 1% | 278 | 0% | 0.337 |
| **Economic Variables** |  |  |  |  |  |  |  |  |  |  |
| Household size | 289 | 4.3 | 290 | 4.4 | 0.597 | 290 | 5.7 | 278 | 5.6 | 0.841 |
| Number of economic dependents | 289 | 3.1 | 290 | 3.1 | 0.941 | 290 | 2.9 | 278 | 3.0 | 0.310 |
| Number that have child economic dependents | 289 | 81% | 290 | 88% | **0.037** | 290 | 82% | 278 | 87% | 0.160 |
| Number of children | 289 | 2.1 | 290 | 1.9 | 0.249 | 290 | 1.2 | 278 | 1.2 | 0.822 |
| Household expenditure per adult equivalent | 289 | 54,804 CFA | 290 | 47,426 CFA | 0.237 | 290 | 32,933 CFA | 278 | 38,776 CFA | 0.101 |
| **Sex work and behaviours** |  |  |  |  |  |  |  |  |  |  |
| Number of clients/ sugar daddies in typical week | 268 | 8.5 | 265 | 8.2 | 0.491 | 290 | 2.8 | 278 | 2.6 | 0.216 |
| Number of sex acts in a typical week | 286 | 17.9 | 288 | 17.4 | 0.578 | 290 | 4.0 | 278 | 3.8 | 0.384 |
| Average earnings from sex work | 289 | 92,332 CFA | 290 | 93,748 CFA | 0.845 | 290 | 45,048 CFA | 278 | 42,986 CFA | 0.748 |
| Used PrEP before | 289 | 9% | 290 | 7% | 0.341 | 290 | 0% | 278 | 0% | 0.308 |
| Had an HIV test in the last 12 months | 288 | 93% | 290 | 92% | 0.893 | 290 | 58% | 277 | 57% | 0.828 |
| Used condom at last sex - direct question | 289 | 85% | 290 | 86% | 0.712 | 290 | 50% | 278 | 50% | 0.867 |
| Used condom at last sex - list experiment | 578 | 0.809 | 580 | 0.798 | #N/ A | 580 | 0.441 | 556 | 0.460 | #N/ A |

### 2.3 Intervention

We designed an insurance product for the purpose of the research in collaboration with the national private insurance company Garantie Mutuelle des Cadres (GMC) Assurances. The insurance product covers the participant and up to six of their nominated economic dependents at randomisation. An economic dependent was defined as anyone who financially depended on the participant to pay health out-of-pocket spending (OOPs) but did not have to be a relative or family member.

Healthcare was freely available to selected participants and covered the costs of consultations, pharmaceutical, medical and hospitalisation fees for non-chronic pathologies from one public district hospital in Yaundé. The Cité Verte Hospital was selected for its advantageous location, fee schedule, capacity, coverage, and cooperation with our intervention. The intervention did not cover COVID-19, chronic diseases, and maternal health care.

The insurance product was implemented as a pre-paid scheme where the hospital recorded and obtained reimbursement from GMC, the insurance company. Participants and their allocated economic dependents had a combined ceiling of health expenses set at CFAF 500,000 (= £644.02 or $776.59)^1^ over the 12 months. The insurance product started on 15^th^ November 2021 and ended on 14^th^ November 2022, with 2,428 people (579 participants allocated to the treatment group and their 1,849 economic dependents) entitled to free healthcare.

To prevent fraudulent use, insurance cards for those eligible to use the health insurance were generated and distributed and needed to be presented to receive free health care alongside an ID document. ID cards were generated by our research team that included a photograph taken on site were distributed to those without valid ID. To minimise stigma at the hospital, women were described as economically vulnerable without mention of being engaged in commercial or transactional sex. Training was conducted with hospital staff to minimise the stigma associated with participants’ characteristics.

### 2.4 Outcomes

The primary outcome measure is HIV infection measured using rapid serological blood tests with positive and undetermined tests re-tested using the enzyme-linked immunosorbent assay (ELISA test). Secondary outcomes measure STIs, including chlamydia, gonorrhoea, trichomoniasis and syphilis. Syphilis was tested using two rapid tests (Venereal Disease Research Laboratory ‘VDRL’ and Treponema Pallidum Haemagglutination ‘VDRL and TPHA’ tests) to determine never-infected, non-active infections (past and treated infections or recent infections) and active infections (non-treated, current infections). All active infections were treated at baseline, so we grouped non-active and never infected together as syphilis-negative. Gonorrhoea and chlamydia were tested using rapid tests, but trichomoniasis was tested in the laboratory as no rapid was available. As per protocols, all positive or inconclusive STI tests from the site were referred for confirmatory tests at the lab, and in all cases, we prioritised the lab results in our analysis (24).

Intermediary outcomes are grouped into behavioural outcomes, including risky behaviours and intensity of commercial or transactional sex; with secondary outcomes of poverty, including expenditure, savings, secondary jobs, and household debt; and secondary mental and physical health outcomes, including self-reliance and depression. This paper only examines the outcomes relating directly to health and risky behaviour outcomes.

We hypothesise the intervention could affect HIV infections through two main channels illustrated in Figure 1. The first is health insurance has been shown to increase healthcare use (27,28), therefore, may lead to an increase in treated STIs, lowering susceptibility to HIV given the existing epidemiological synergy between STIs and HIV (29). However, ex-ante moral hazard may lead to an increase in risky behaviours as a result of receiving the intervention that could mitigate any beneficial impacts. The second channel is through the reduction in health shocks resulting from the elimination of OOPs that should prevent the need to engage in risky sexual behaviours (17,30). Specifically, an increased probability of leaving the risky sex market (extensive margin outcome), a decrease in the number of sex acts and sexual partners (intensive margin outcomes) or a reduction in the likelihood of engaging in risky sex (e.g. unprotected sex and anal sex) that will lead to a reduction in the incidence of STIs and HIV.

**Figure 1:**
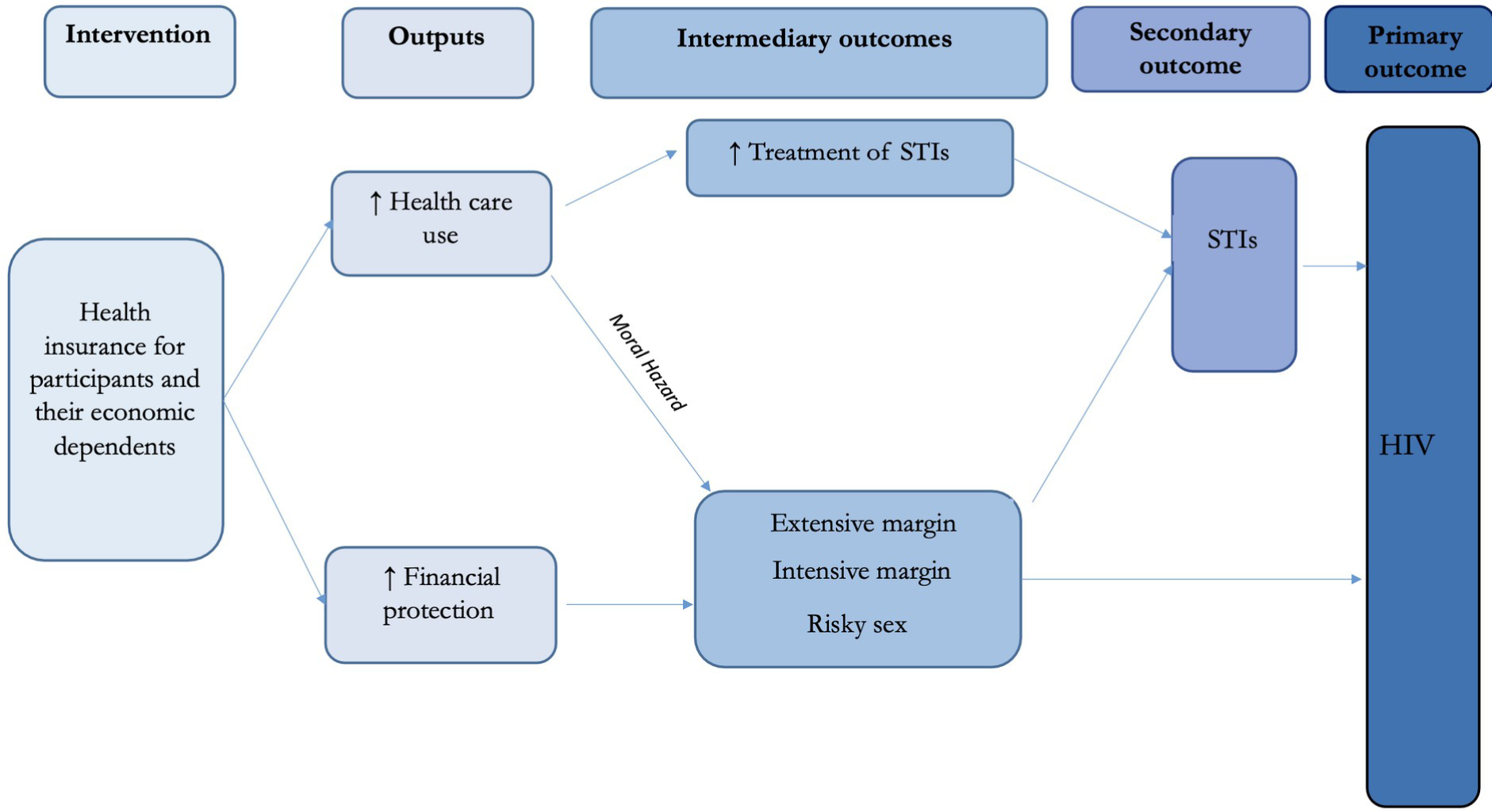
Theory of change

It is worth noting that risky sexual behaviours such as unprotected sex are prone to sensitivity bias (31,32). To minimise social desirability bias, we ensured the privacy of interviews and recruited enumerators from the same community and population as the study participants and we collected unprotected sex using the double-list experiment method (See appendix 7.2 for details).

### 2.5 Sample size and attrition

Power calculations were performed to determine the required sample size and detectable minimum effects (MDE). It was assumed that 600 participants out of the 750 recruited at baseline would be retained at the endline, and assuming that 20% of participants would acquire at least one STI over the trial period, the sample size was sufficient to detect a decrease of 30% (6 percentage points) in the incidence of STIs in the treatment group assuming 80% power and a significance level of 5%. To conduct analysis separately among the sub-samples of those engaging in commercial and transactional sex, the MDE is a ∼40% reduction (8.3 percentage points) in STI incidence. Assuming an HIV incidence of 4% over the trial period, the sample size would allow the detection of a decrease of 65% in HIV incidence (2.6 percentage points).

Those assumptions were close to the number we retained within the trial. A total of 956 (493 transactional and 463 commercial) respondents completed the survey and biological tests at midline, and 806 (403 transactional and 403 commercial) did so for endline. Combined 1,020 (530 transactional and 490 commercial) respondents completed the survey and biological tests at midline or endline, indicating that one post-intervention survey was available for 89% of the randomised sample.

In Table 2, we see the response rates of each arm and strata of the study. Attrition by endline was around 30%. We do find that attritors are younger and more likely to be part of the commercial sex arm. In the transactional sex arm, attritors are more likely to be part of the treatment group. However, we find no statistically significant difference between the attritors in the treatment and control arms of the survey in commercial, transactional and pooled strata, Tables Table A4 and Table A5. There were no unintended effect or harm reported as a result of this intervention.

**Table 2:** Response rates.

|  | Pooled | Transactional | Commercial |
| --- | --- | --- | --- |
| Baseline & Randomisation | 1146 (100%) | 568 (100%) | 578 (100%) |
| Midline | 956 (83%) | 493 (87%) | 463 (80%) |
| Endline | 806 (70%) | 403 (71%) | 403 (70%) |
| Midline or Endline | 1020 (89%) | 530 (93%) | 490 (85%) |

Our final analysis sample comprises endline surveys plus any midline surveys and STI test results for those that did not complete endline surveys, see row 4 of Table 2. This analysis sample is the one we use throughout the paper, and all adjusted models include an adjustment for results that were collected at midline since they only had six months of treatment versus the entire year for those with endline results.

### 2.6 Statistical Analysis

Data analysis was blinded to the allocation of respondents. All analyses have been conducted in STATA version 17 and follow the analysis specified in the study protocol (24) and registered on ISRCTN (study 225165484). To assess the effect of health insurance on our primary and secondary outcomes, we conducted an intention-to-treat (ITT) analysis where respondents were analysed based on their randomised study group. We estimate logistic regressions with odds ratios for our primary biological binary outcomes. We report the marginal effect for all other binary outcomes. We estimate the marginal effects using ordinary least squares (OLS) for continuous outcomes. Table notes indicate which outcomes were estimated via which method. Adjusted models control for outcome level at baseline, logged adult equivalent total expenditure as a proxy for income, the number of economic dependents, the household size and if the respondent has a child.

## 3 Results

A total of 938 medical visits were conducted over 12 months, including 411 participant patients (44%) and 527 economic dependent patients (56.1%). The main reasons for consultations included malaria (34%), STI treatment based on syndromic case management (14%), cold and flu (14%), gastrointestinal disease (11%), skin problems (5%), lung infection (8%), pelvic pain (7%), accident and injuries (3%), see Figure A1. For participants (excluding economic dependents), there were 123 consultations (30%) for STIs.

### 3.1 Effect of Health Insurance on HIV and other STIs

We find the intervention to be effective to prevent HIV. The intervention significantly reduces the odds of acquiring HIV for the transactional sex strata (OR=0.109, p<0.05; AOR = 0.113, p<0.05) and when the samples are pooled (OR=0.197, p<0.05; AOR = 0.204, p<0.05), see Table A0 with odd ratio model estimates in Table A0 with marginal effects reported in Table A8. There is no impact on HIV for women engaging in commercial sex, but as shown in Table A2, the number of HIV infections is low over the period among this sub-group.

We find no effect of the intervention for other STIs, see Table A0. The absence of effect could be due to serious issues found with the STI tests because they were not effective in determining certainty the STI status. Every 15^th^ chlamydia, syphilis and HIV test performed, plus any positive and inconclusive test, was verified in labs using the same rapid tests. At endline, we verified as many samples as possible were re-tested at the ELISA laboratory with new types of tests. Table 3 shows large type-II errors in detecting chlamydia, imprecision in detecting syphilis, but consistent results for HIV. There is evidence that the tests for chlamydia and syphilis lacked sensitivity and that there were issues with sample acquisition, transportation, and storage for gonorrhoea and trichomoniasis tests in the field.

**Table 3:** Result of rapid test vs. laboratory ELISA tests.

|  | Rapid test | ELISA laboratory test |
| --- | --- | --- |
| **HIV** |  |  |
| Positive | 5 | 5 |
| Negative | 57 | 58 |
| Inconclusive | 1 | 0 |
| **Chlamydia** |  |  |
| Positive | 1 | 57 |
| Negative | 202 | 149 |
| Inconclusive | 11 | 8 |
| **Syphilis** |  |  |
| Positive active infection | 2 | 0 |
| Negative | 49 | 63 |
| Positive - inactive infection | 0 | 11 |
| Inconclusive | 23 | 0 |

Note: only endline samples that had both a rapid and ELISA tests are included.

Appendix 7.3 contains further details.

### 3.2 Effect on risky sexual behaviours

Table 4 shows the impact of the intervention on risky sexual behaviours. The intervention led to a significant increase in the likelihood of quitting transactional sex by 8.6 percentage points (50% increase) in likelihood of leaving sex work. There is an increase in condom use at the last sex act of around 15 percentage points, or a 43% increase, when measured using the list experiment (ME=0.152) and we observe a decrease in the number of sex acts by 0.36 on average for women in transactional sex (16% drop), which are statistically significance at 10%. For commercial sex, health insurance had mixed results on risky behaviours. The intervention was not effective in allowing women to leave sex work and did not increase condom use among sex workers but led to small increases in anal sex (ME=0.014) and riskiness of clients (ME=0.039). Only the small increase in the riskiness of clients / sugar daddies is maintained when pooled, suggesting highly differential behaviour impacts of health insurance on women in commercial and transactional sex. We report unadjusted and adjusted odd ratios in Appendix 7.1 in Table A7.

**Table 4:** Effect of health insurance on sexual behaviours behaviour outcomes.

| Sexual Behaviour Outcomes | Unadjusted |  |  | Adjusted |  |  |
| --- | --- | --- | --- | --- | --- | --- |
|  | n | ME | 95% CI | n | ME | 95% CI |
| **Commercial Sex** |  |  |  |  |  |  |
| Number of regular clients in the last 7 days | 363 | 0.125 | -0.381 - 0.631 | 363 | 0.098 | -0.409 - 0.606 |
| Number of occasional clients in the last 7 days | 355 | 0.340 | -0.226 - 0.906 | 355 | 0.274 | -0.290 - 0.838 |
| Number of clients in the last 7 days (combined) | 489 | 0.199 | -0.520 - 0.919 | 489 | 0.151 | -0.550 - 0.853 |
| Number of sex acts with clients in the last 7 days | 489 | -0.242 | -1.352 - 0.868 | 489 | -0.364 | -1.447 - 0.718 |
| Number of vaginal sex acts‡ | 730 | 0.002 | -0.014 - 0.018 | 730 | 0.002 | -0.014 - 0.018 |
| Number of oral sex acts‡ | 729 | -0.046* | -0.093 - 0.001 | 729 | -0.036 | -0.082 - 0.010 |
| Number of anal sex acts‡ | 730 | 0.014** | 0.002 - 0.026 | 730 | 0.013** | 0.001 - 0.025 |
| Self-reported riskiness of clients (average / 10)‡ | 730 | 0.039** | 0.009 - 0.070 | 730 | 0.038** | 0.007 - 0.068 |
| Used condom at last sex act - Direct questioning† | 364 | 0.014 | -0.051 - 0.079 | 364 | 0.013 | -0.051 - 0.077 |
| Quit commercial sex work† | 439 | 0.008 | -0.070 - 0.085 | 439 | 0.008 | -0.067 - 0.084 |
| **List Experiment** |  |  |  |  |  |  |
| Used condom at last sex act - List experiment | 730 | -0.111 | -0.348 - 0.125 | 730 | -0.109 | -0.345 - 0.128 |
| **Transactional Sex** |  |  |  |  |  |  |
| Number of regular clients in the last 7 days | 399 | 0.059 | -0.190 - 0.307 | 399 | 0.080 | -0.169 - 0.330 |
| Number of occasional clients in the last 7 days | 399 | -0.021 | -0.191 - 0.150 | 399 | 0.003 | -0.170 - 0.176 |
| Number of papy's in the last 7 days | 530 | -0.164 | -0.439 - 0.110 | 530 | -0.140 | -0.414 - 0.133 |
| Number of sex acts in the last 7 days | 530 | -0.358* | -0.781 - 0.065 | 530 | -0.320 | -0.742 - 0.102 |
| Number of vaginal sex acts‡ | 756 | 0.006 | -0.011 - 0.024 | 756 | 0.007 | -0.011 - 0.024 |
| Number of oral sex acts‡ | 757 | -0.023 | -0.074 - 0.027 | 757 | -0.019 | -0.068 - 0.030 |
| Number of anal sex acts‡ | 758 | 0.001 | -0.012 - 0.014 | 758 | -0.001 | -0.014 - 0.012 |
| Self-reported riskiness of clients (average / 10)‡ | 758 | 0.014 | -0.012 - 0.039 | 758 | 0.008 | -0.018 - 0.034 |
| Used condom at last sex act - Direct questioning† | 399 | 0.023 | -0.074 - 0.120 | 399 | 0.016 | -0.082 - 0.114 |
| Quit transactional sex work† | 496 | 0.086** | 0.015 - 0.158 | 496 | 0.084** | 0.013 - 0.155 |
| **List Experiment** |  |  |  |  |  |  |
| Used condom at last sex act - List experiment | 798 | 0.152* | -0.021 - 0.324 | 798 | 0.150* | -0.025 - 0.325 |
| **Pooled (commercial + transactional)** |  |  |  |  |  |  |
| **Continuous outcomes - OLS** |  |  |  |  |  |  |
| Number of regular clients in the last 7 days | 762 | 0.135 | -0.158 - 0.428 | 762 | 0.092 | -0.191 - 0.376 |
| Number of occasional clients in the last 7 days | 754 | 0.233 | -0.077 - 0.544 | 754 | 0.167 | -0.132 - 0.466 |
| Number of papy's or clients in the last 7 days | 1,019 | 0.030 | -0.398 - 0.457 | 1,019 | -0.010 | -0.418 - 0.397 |
| Number of sex acts in the last 7 days | 1,019 | -0.279 | -0.891 - 0.333 | 1,019 | -0.391 | -0.969 - 0.187 |
| Number of vaginal sex acts‡ | 1,486 | 0.004 | -0.007 - 0.016 | 1,486 | 0.005 | -0.007 - 0.017 |
| Number of oral sex acts‡ | 1,486 | -0.035** | -0.069 - -0.001 | 1,486 | -0.024 | -0.058 - 0.009 |
| Number of anal sex acts‡ | 1,488 | 0.007 | -0.001 - 0.016 | 1,488 | 0.007 | -0.002 - 0.016 |
| Self-reported riskiness of clients (average / 10)‡ | 1,488 | 0.031*** | 0.008 - 0.054 | 1,488 | 0.027** | 0.005 - 0.049 |
| Used condom at last sex act - Direct questioning† | 763 | 0.014 | -0.051 - 0.079 | 763 | 0.036 | -0.029 - 0.101 |
| Quit either commercial or transactional† | 935 | 0.008 | -0.070 - 0.085 | 935 | 0.043 | -0.008 - 0.095 |
| **List Experiment** |  |  |  |  |  |  |
| Used condom at last sex act - List experiment | 1,528 | 0.034 | -0.113 - 0.180 | 1,528 | 0.041 | -0.106 - 0.187 |

Notes: Dependent binary variables (†) were estimated using logistic regression reporting marginal effects. All other continuous outcome variables were estimated using OLS reporting marginal effects. (log) indicate where the outcome is logged in the OLS model. Unadjusted = compares outcomes of follow-up data without adjustment. Adjusted = adjusts for baseline levels of the outcome, if the outcome was collected during midline, then baseline controls, namely log of adult equivalent expenditure, number of economic dependents, household size, if the respondent has a child and if the respondent is enrolled in school. The list experiment was estimated using OLS using the double list experiment method (Lépine et al., 2020) hence twice the observations as each respondent appears twice.
‡= Data taken from the last two sex acts per sex worker.

### 3.3 Analysis of the intervention channels and alternative explanations

A key question regarding this intervention is the pathway and mechanism in which it operates. As per Figure 1, its impact on HIV can occur through two potential channels. 1) by increasing treatment of STIs, lowers the biological susceptibility to HIV when engaging in unprotected sex, and 2) reducing the prevalence and associated risky behaviours from economic shocks. To disentangle these two channels, we investigate the impact of the intervention on STIs and economic shocks.

First, we can examine the role of STI as measured by self-reported STI symptoms in mediating the intervention’s impact on HIV in the transactional arm. Note that utilisation of healthcare is strongly correlated with protection against health expense shocks, and test results for STIs are questionable. Symptoms of STIs that increase HIV susceptibility are typically visible or noticeable to the individual, whereas HIV symptoms are not and can remain hidden for years after the infection. Of women engaged in transactional sex, ten had acquired HIV by endline, nine in the control group and one in the treatment group, see Table A2. 17 women reported STI symptoms^2^, with 13 of these in the control group^3^. After discounting HIV infections among people who also reported STI symptoms, we see that the only participant in the treatment group who was HIV positive also reported STI symptoms between baseline and endline while only one participant of the control group who was HIV positive reported STI symptoms. This leaves eight HIV infections without association with STI symptoms.

In addition, Table A3 displays results from models with self-reported STI symptoms as an interaction term, finding the impact of the intervention persists in reducing HIV by around 3.5 percentage points both when conditioning on STI symptoms (ME = −0.036, p<0.05) and when removing those with STI symptoms (ME = −0.034, p<0.05). There is only a low incidence of reported STI symptoms, so we treat these estimates cautiously, but they suggest the intervention is not acting through the STI treatment channel.

We now investigate the role of reduced economic shocks. On average, 70% of insured participants or their economic dependents were sick at least once over the treatment period. 57% of participants reported receiving free care due to the intervention for themselves or their economic dependents. Table 5 shows the reduction in expenses and health shocks for the transactional sex group. Binary outcomes also have unadjusted and adjusted odd ratios reported in Table A6. We cannot perform further statistical analysis conditioning on utilisation or shock protection since the intervention will change behaviours in anticipation of not having health costs should they arise. Overall, however, this evidence suggests the channel through which the intervention works is the protection against economic shocks.

**Table 5:**
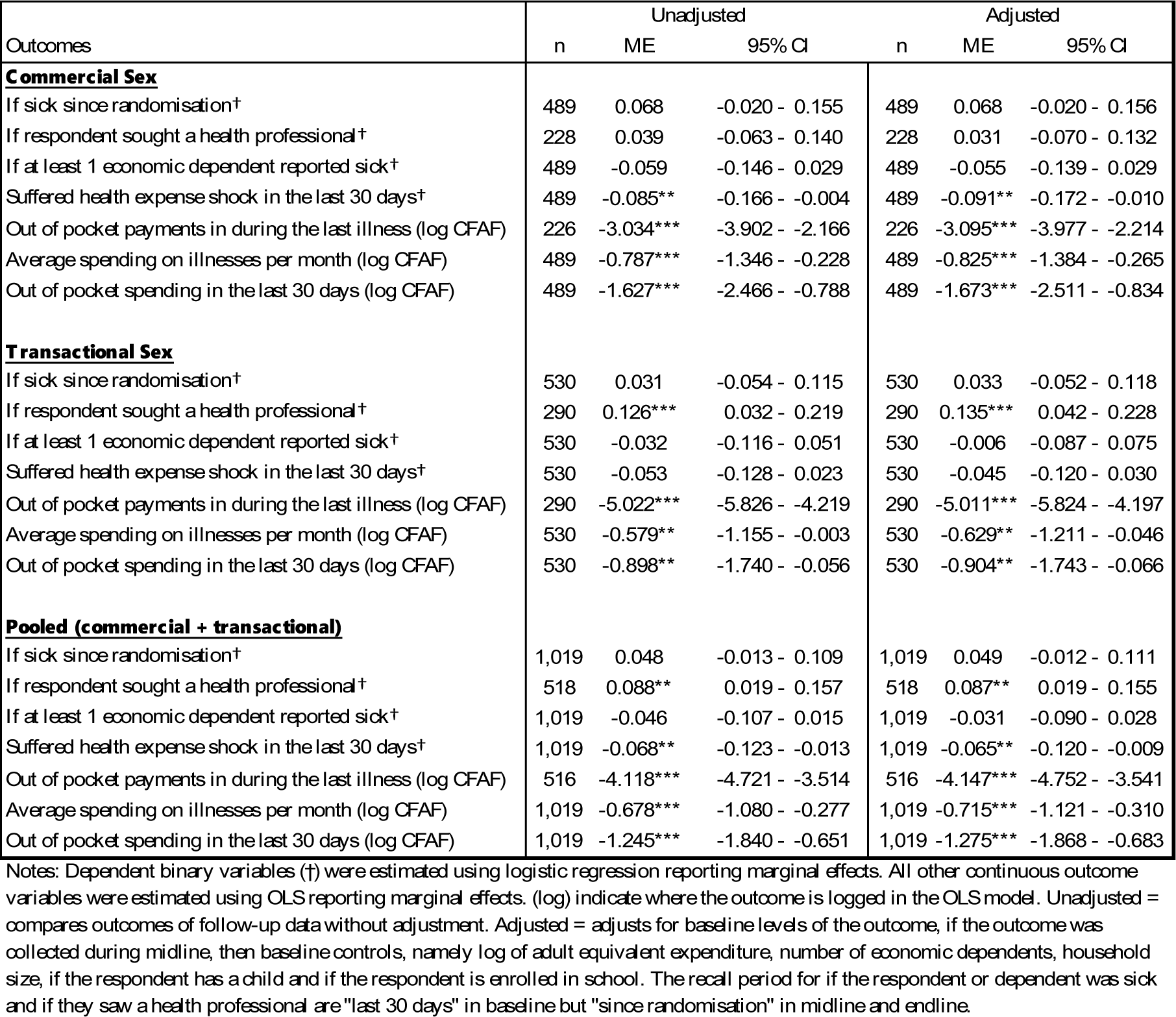
Effect of health insurance on health utilisation and health spending outcomes.

| Outcomes | Unadjusted |  |  | Adjusted |  |  |
| --- | --- | --- | --- | --- | --- | --- |
|  | n | ME | 95% CI | n | ME | 95% CI |
| **Commercial Sex** |  |  |  |  |  |  |
| If sick since randomisation† | 489 | 0.068 | -0.020 - 0.155 | 489 | 0.068 | -0.020 - 0.156 |
| If respondent sought a health professional† | 228 | 0.039 | -0.063 - 0.140 | 228 | 0.031 | -0.070 - 0.132 |
| If at least 1 economic dependent reported sick† | 489 | -0.059 | -0.146 - 0.029 | 489 | -0.055 | -0.139 - 0.029 |
| Suffered health expense shock in the last 30 days† | 489 | -0.085** | -0.166 - -0.004 | 489 | -0.091** | -0.172 - -0.010 |
| Out of pocket payments in during the last illness (log CFAF) | 226 | -3.034*** | -3.902 - -2.166 | 226 | -3.095*** | -3.977 - -2.214 |
| Average spending on illnesses per month (log CFAF) | 489 | -0.787*** | -1.346 - -0.228 | 489 | -0.825*** | -1.384 - -0.265 |
| Out of pocket spending in the last 30 days (log CFAF) | 489 | -1.627*** | -2.466 - -0.788 | 489 | -1.673*** | -2.511 - -0.834 |
| **Transactional Sex** |  |  |  |  |  |  |
| If sick since randomisation† | 530 | 0.031 | -0.054 - 0.115 | 530 | 0.033 | -0.052 - 0.118 |
| If respondent sought a health professional† | 290 | 0.126*** | 0.032 - 0.219 | 290 | 0.135*** | 0.042 - 0.228 |
| If at least 1 economic dependent reported sick† | 530 | -0.032 | -0.116 - 0.051 | 530 | -0.006 | -0.087 - 0.075 |
| Suffered health expense shock in the last 30 days† | 530 | -0.053 | -0.128 - 0.023 | 530 | -0.045 | -0.120 - 0.030 |
| Out of pocket payments in during the last illness (log CFAF) | 290 | -5.022*** | -5.826 - -4.219 | 290 | -5.011*** | -5.824 - -4.197 |
| Average spending on illnesses per month (log CFAF) | 530 | -0.579** | -1.155 - -0.003 | 530 | -0.629** | -1.211 - -0.046 |
| Out of pocket spending in the last 30 days (log CFAF) | 530 | -0.898** | -1.740 - -0.056 | 530 | -0.904** | -1.743 - -0.066 |
| **Pooled (commercial + transactional)** |  |  |  |  |  |  |
| If sick since randomisation† | 1,019 | 0.048 | -0.013 - 0.109 | 1,019 | 0.049 | -0.012 - 0.111 |
| If respondent sought a health professional† | 518 | 0.088** | 0.019 - 0.157 | 518 | 0.087** | 0.019 - 0.155 |
| If at least 1 economic dependent reported sick† | 1,019 | -0.046 | -0.107 - 0.015 | 1,019 | -0.031 | -0.090 - 0.028 |
| Suffered health expense shock in the last 30 days† | 1,019 | -0.068** | -0.123 - -0.013 | 1,019 | -0.065** | -0.120 - -0.009 |
| Out of pocket payments in during the last illness (log CFAF) | 516 | -4.118*** | -4.721 - -3.514 | 516 | -4.147*** | -4.752 - -3.541 |
| Average spending on illnesses per month (log CFAF) | 1,019 | -0.678*** | -1.080 - -0.277 | 1,019 | -0.715*** | -1.121 - -0.310 |
| Out of pocket spending in the last 30 days (log CFAF) | 1,019 | -1.245*** | -1.840 - -0.651 | 1,019 | -1.275*** | -1.868 - -0.683 |

Notes: Dependent binary variables (†) were estimated using logistic regression reporting marginal effects. All other continuous outcome variables were estimated using OLS reporting marginal effects. (log) indicate where the outcome is logged in the OLS model. Unadjusted = compares outcomes of follow-up data without adjustment. Adjusted = adjusts for baseline levels of the outcome, if the outcome was collected during midline, then baseline controls, namely log of adult equivalent expenditure, number of economic dependents, household size, if the respondent has a child and if the respondent is enrolled in school. The recall period for if the respondent or dependent was sick and if they saw a health professional are "last 30 days" in baseline but "since randomisation" in midline and endline.

In Appendix 7.5, we also present evidence that our results are not driven by ex-ante moral hazard since the intervention does not increase the likelihood to report an illness, nor an increase in risky health behaviours.

## 4 Discussion

We find that health insurance provides protection against economic shocks for all and protection against HIV acquisition for women engaged in transactional sex. We find strong evidence that the primary mechanism through which the intervention works is by protecting against health expense shocks that reduce risky sexual behaviours. Our findings are robust to removing HIV acquired through increased susceptibility because of reported STI symptoms.

Women engaged in commercial sex did not benefit in the same way as women engaged in transactional sex, and this could be due to the additional attention, resources and protections afforded to them as a “key population” in fighting HIV (2). Specifically, FSWs have greater access to PrEP and other educational resources on HIV. 0% of women in transactional sex had taken PrEP before, compared to just under 10% of women in commercial sex. Also, over 90% of women in commercial sex had been tested for HIV in the last 12 months, compared to only 50% of women in transactional sex, see Table 1. On the other hand, transactional relationships are built on the basis of trust, which could explain the observed lower condom use on average among this arm of the study.

A limitation of our primary HIV findings is the small number of positive infections over the 12 month period (2.0% in the control group) which is lower than the official statistics of 4% estimated in Cameroon (24). As a robustness check, we used bootstrapping techniques over 1000 resamples, finding similar results, see Table A2. The suspected issues with chlamydia and syphilis are important and worrying since the tests used in this study are the Cameroon Health Authority’s recommended tests. Should they be inaccurate, it could indicate significant country-level type-II error with major implications for the national HIV prevention strategy.

This is the first study to highlight the critical heterogeneity of women who engage in transactional sex and female sex workers in the study of HIV prevention. Previous literature has often conflated commercial and transactional sex (5). The more recent debate focuses on the similarities in their structural drivers (33,34) and questions the usefulness of having distinct categories (35). However, this study quantitatively highlights the differential risks taken by women engaging in commercial and transactional sex, meaning HIV prevention needs to be aware of the nuanced differences. Future research should consider some degree of categorisation to understand the differential impacts of HIV prevention interventions among these separate groups.

Our results show the importance of transactional sex in HIV transmission since we find 9 times more infections among women in transactional sex than in women in commercial sex in the control group. We contribute to the growing evidence that women engaging in transactional sex should be considered as a key population in the fight against HIV. We show how a structural intervention is more protective against HIV for women in transactional sex than FSWs, yet currently they receive far fewer HIV prevention services and are ineligible to PrEP (2).

We perform a back-of-the-envelope calculation for the cost-effectiveness. The total cost of healthcare claimed by the intervention was £42,003, which averted 9 HIV infections in the transactional sex arm. This gives a cost-effectiveness ratio per HIV infection averted of £4,667 for women engaged in transactional sex. This means the intervention would likely be highly cost-effective when scaled up, and this figure does not account for the positive externalities involved in the prevention of HIV in broader society.

To conclude, our study provides the first evidence of an intervention directly targeting health expense shocks, the most common economic shock to affect women engaging in commercial and transactional sex in urban environments, as a strategy to prevent HIV among women. Our study highlights the importance of including women engaging in transactional sex in Africa as a key population to receive additional support and protection in the fight against HIV.

## Data Availability

All relevant data are within the manuscript and its Supporting Information files.

## 5 Funding

We acknowledge funding from UKRI.

### 7 Appendix

#### 7.1 Additional Results Tables and Figures

**Figure A0:**
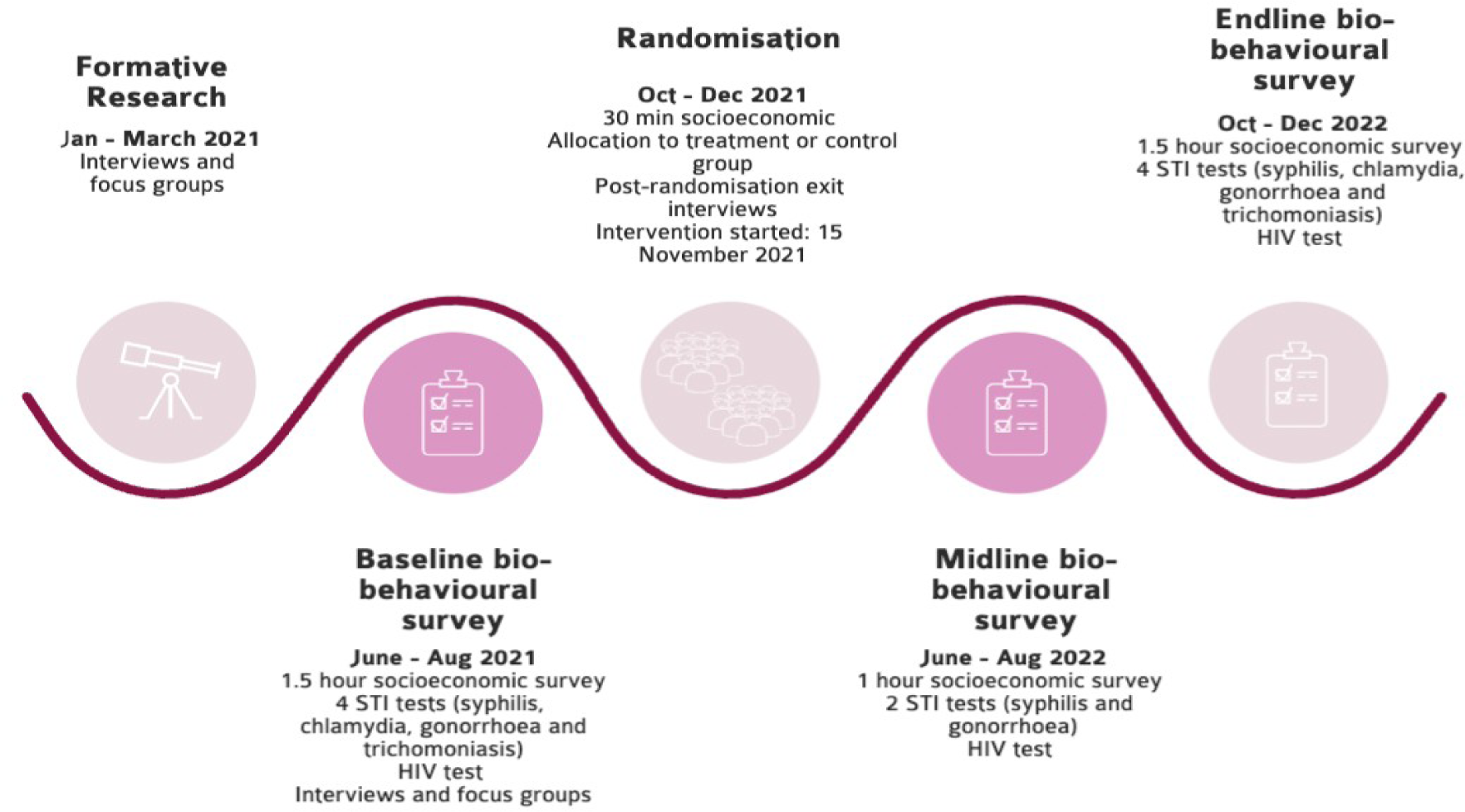
Project timeline

**Figure A1:**
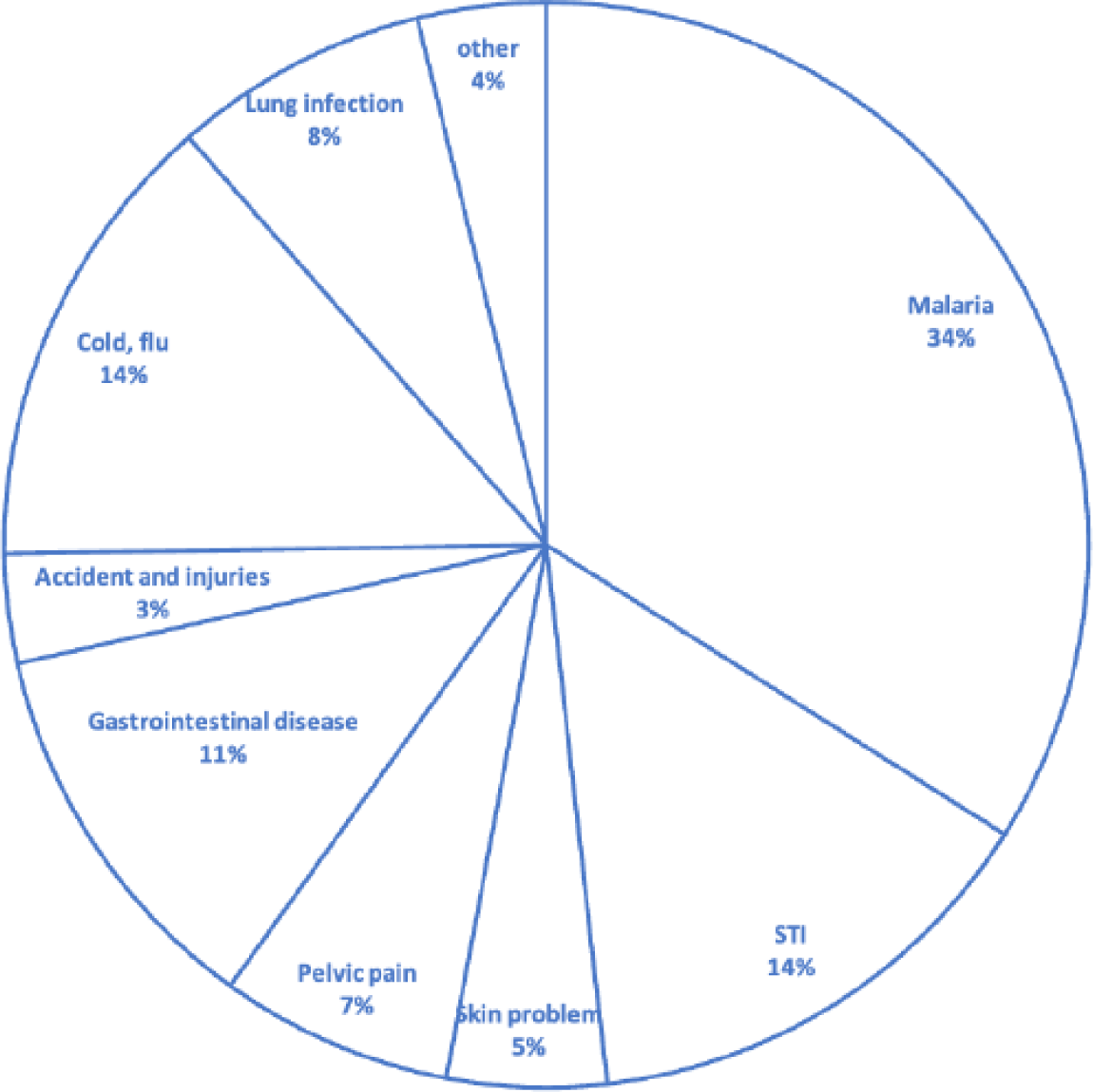
Reason for consultation

**Table A0:** Effect of health insurance on biological outcomes.

| Outcomes | Baseline |  |  |  |  |  | Midline/ Endline |  |  |  |  |  | Difference |  |  |  |
| --- | --- | --- | --- | --- | --- | --- | --- | --- | --- | --- | --- | --- | --- | --- | --- | --- |
|  | Magenta |  |  | Teal |  |  | Magenta |  |  | Teal |  |  | Unadjusted odds |  | Adjusted odds |  |
|  | obs | n | %/ mean | obs | n | %/ mean | obs | n | %/ mean | obs | n | %/ mean | OR | 95% CI | AOR | 95% CI |
| **Commercial Sex** |  |  |  |  |  |  |  |  |  |  |  |  |  |  |  |  |
| **Biological** |  |  |  |  |  |  |  |  |  |  |  |  |  |  |  |  |
| HIV | 289 | 0 | 0.0% | 289 | 0 | 0.0% | 246 | 1 | 0.4% | 243 | 1 | 0.4% | 0.988 | 0.061 - 15.882 | 0.854 | 0.046 - 15.958 |
| HIV - bootstrapped | - | - | - | - | - | - | - | - | - | - | - | - | 0.988 | 0.270 - 3.616 | 0.854 | 0.241 - 3.035 |
| Syphilis (active) | 287 | 23 | 8.0% | 288 | 23 | 8.0% | 246 | 1 | 0.4% | 243 | 4 | 1.6% | 0.244 | 0.027 - 2.198 | 0.526 | 0.045 - 6.146 |
| Gonorrhoea | 230 | 30 | 13.0% | 246 | 28 | 11.4% | 231 | 1 | 0.4% | 227 | 1 | 0.4% | 0.983 | 0.061 - 15.805 | 0.728 | 0.022 - 23.798 |
| Chlamydia | 286 | 3 | 1.0% | 288 | 2 | 0.7% | 201 | 15 | 7.5% | 201 | 12 | 6.0% | 1.270 | 0.579 - 2.786 | 1.310 | 0.586 - 2.926 |
| Trichomoniasis | 230 | 2 | 0.9% | 246 | 4 | 1.6% | 178 | 1 | 0.6% | 182 | 3 | 1.6% | 0.337 | 0.035 - 3.272 | 0.334 | 0.032 - 3.533 |
| **Transactional Sex** |  |  |  |  |  |  |  |  |  |  |  |  |  |  |  |  |
| **Biological** |  |  |  |  |  |  |  |  |  |  |  |  |  |  |  |  |
| HIV | 290 | 0 | 0.0% | 278 | 0 | 0.0% | 263 | 1 | 0.4% | 267 | 9 | 3.4% | 0.109** | 0.014 - 0.870 | 0.113** | 0.014 - 0.912 |
| HIV - bootstrapped | - | - | - | - | - | - | - | - | - | - | - | - | 0.109*** | 0.036 - 0.336 | 0.113*** | 0.037 - 0.345 |
| Syphilis (active) | 289 | 0 | 0.0% | 278 | 1 | 0.4% | 263 | 10 | 3.8% | 267 | 3 | 1.1% | 3.478* | 0.946 - 12.784 | 3.136 | 0.786 - 12.506 |
| Gonorrhoea | 256 | 17 | 6.6% | 250 | 11 | 4.4% | 262 | 7 | 2.7% | 267 | 4 | 1.5% | 1.805 | 0.522 - 6.240 | 1.524 | 0.391 - 5.945 |
| Chlamydia | 289 | 2 | 0.7% | 278 | 1 | 0.4% | 192 | 16 | 8.3% | 209 | 15 | 7.2% | 1.176 | 0.565 - 2.448 | 1.171 | 0.556 - 2.464 |
| Trichomoniasis | 256 | 5 | 2.0% | 250 | 5 | 2.0% | 192 | 7 | 3.6% | 210 | 6 | 2.9% | 1.286 | 0.425 - 3.898 | 1.359 | 0.443 - 4.165 |
| **Pooled (commercial + transactional)** |  |  |  |  |  |  |  |  |  |  |  |  |  |  |  |  |
| **Biological** |  |  |  |  |  |  |  |  |  |  |  |  |  |  |  |  |
| HIV | 579 | 0 | 0.0% | 567 | 0 | 0.0% | 509 | 2 | 0.4% | 510 | 10 | 2.0% | 0.197** | 0.043 - 0.905 | 0.204** | 0.044 - 0.940 |
| HIV - bootstrapped | - | - | - | - | - | - | - | - | - | - | - | - | 0.197** | 0.057 - 0.680 | 0.204** | 0.054 - 0.769 |
| Syphilis (active) | 576 | 23 | 4.0% | 566 | 24 | 4.2% | 509 | 11 | 2.2% | 510 | 7 | 1.4% | 1.587 | 0.610 - 4.127 | 1.972 | 0.676 - 5.753 |
| Gonorrhoea | 486 | 47 | 9.7% | 496 | 39 | 7.9% | 493 | 8 | 1.6% | 494 | 5 | 1.0% | 1.613 | 0.524 - 4.966 | 1.511 | 0.450 - 5.075 |
| Chlamydia | 575 | 5 | 0.9% | 566 | 3 | 0.5% | 393 | 31 | 7.9% | 410 | 27 | 6.6% | 1.215 | 0.711 - 2.075 | 1.215 | 0.707 - 2.090 |
| Trichomoniasis | 486 | 7 | 1.4% | 496 | 9 | 1.8% | 370 | 8 | 2.2% | 392 | 9 | 2.3% | 0.940 | 0.359 - 2.464 | 1.007 | 0.383 - 2.648 |

Notes: Unadjusted = logistic regression of follow-up data. Adjusted = Logistic regression adjusting treatment effect for baseline levels of outcomes, if the outcome was collected during midline, then baseline controls, namely log of adult equivalent expenditure, number of economic dependents, household size, if the respondent has a child and if the respondent is enrolled in school. Trichomoniasis, Vaginalis, Mobiluncus and Candida were not tested at baseline. The intervention was distributed to HIV negative respondents only.

**Table A1:** Effect of health insurance on moral hazard.

| Sexual Behaviour Outcomes | Unadjusted |  |  | Adjusted |  |  |
| --- | --- | --- | --- | --- | --- | --- |
|  | n | ME | 95% CI | n | ME | 95% CI |
| **Commercial Sex** |  |  |  |  |  |  |
| Number of vaginal sex acts‡ | 140 | -0.005 | -0.046 0.035 | 140 | -0.005 | -0.047 0.036 |
| Number of oral sex acts‡ | 140 | -0.085 | -0.201 0.032 | 140 | -0.061 | -0.175 0.053 |
| Number of anal sex acts‡ | 140 | 0.034* | -0.006 0.075 | 140 | 0.031 | -0.010 0.072 |
| Self-reported riskiness of clients (average / 10)‡ | 140 | 0.033 | -0.031 0.096 | 140 | 0.057* | -0.005 0.119 |
| Used condom at last sex act - Direct questioning† | 121 | 0.066 | -0.057 0.190 | 121 | 0.069 | -0.056 0.194 |
| **List Experiment** |  |  |  |  |  |  |
| Used condom at last sex act - List experiment | 242 | -0.110 | -0.530 0.309 | 242 | -0.103 | -0.536 0.330 |
| **Transactional Sex** |  |  |  |  |  |  |
| Number of vaginal sex acts‡ | 152 | -0.001 | -0.037 0.036 | 152 | 0.001 | -0.037 0.040 |
| Number of oral sex acts‡ | 152 | -0.084 | -0.199 0.031 | 152 | -0.103* | -0.218 0.011 |
| Number of anal sex acts‡ | 152 | 0.000 | 0.000 0.000 | 152 | 0.000 | 0.000 0.000 |
| Self-reported riskiness of clients (average / 10)‡ | 152 | 0.022 | -0.031 0.074 | 152 | 0.017 | -0.037 0.072 |
| Used condom at last sex act - Direct questioning† | 109 | 0.196** | 0.031 0.361 | 109 | 0.185** | 0.021 0.349 |
| **List Experiment** |  |  |  |  |  |  |
| Used condom at last sex act - List experiment | 218 | 0.180 | -0.165 0.524 | 218 | 0.158 | -0.205 0.522 |
| **Pooled (commercial + transactional)** |  |  |  |  |  |  |
| **Continuous outcomes - OLS** |  |  |  |  |  |  |
| Number of vaginal sex acts‡ | 292 | -0.003 | -0.030 0.024 | 292 | -0.001 | -0.028 0.027 |
| Number of oral sex acts‡ | 292 | -0.083** | -0.164 -0.002 | 292 | -0.069* | -0.150 0.011 |
| Number of anal sex acts‡ | 292 | 0.015 | -0.004 0.034 | 292 | 0.015 | -0.004 0.034 |
| Self-reported riskiness of clients (average / 10)‡ | 292 | 0.017 | -0.027 0.060 | 292 | 0.021 | -0.022 0.065 |
| Used condom at last sex act - Direct questioning† | 230 | 0.102* | -0.019 0.224 | 230 | 0.129** | 0.009 0.249 |
| **List Experiment** |  |  |  |  |  |  |
| Used condom at last sex act - List experiment | 460 | 0.013 | -0.265 0.291 | 460 | 0.019 | -0.265 0.303 |

Notes: Dependent binary variables (†) were estimated using logistic regression reporting marginal effects. Continuous outcome variables were estimated using OLS reporting marginal effects. (log) indicate where the outcome is logged in the OLS model. Unadjusted = compares outcomes of follow-up data without adjustment. Adjusted = adjusts for baseline levels of the outcome, if the outcome was collected during midline, then baseline controls, namely log of adult equivalent expenditure, number of economic dependents, household size, if the respondent has a child and if the respondent is enrolled in school. The list experiment was estimated using OLS using the double list experiment method (Lépine et al., 2020) hence twice the observations as each respondent appears twice. ‡ = Data taken from the last two sex acts per sex worker.

**Table A2:** HIV and self-reported STI symptoms for women in the transactional sex arm.

|  | Treatment |  | Control |  |
| --- | --- | --- | --- | --- |
|  | HIV positive | HIV negative | HIV positive | HIV negative |
| HIV Incidence between baseline and follow-up | 1 | 262 | 9 | 258 |
| Self-reported STI or UTI symptoms | 1 | 3 | 1 | 12 |
| HIV infections without STI/ UTI symptoms reported | 0 | 259 | 8 | 246 |

**Table A3:** Effect of health insurance on HIV for those engaged in transactional sex accounting for STI symptoms.

|  | Unadjusted odds |  |  | Adjusted odds |  |  |
| --- | --- | --- | --- | --- | --- | --- |
|  | n | ME | 95% CI | n | AME | 95% CI |
| **Transactional Sex** |  |  |  |  |  |  |
| HIV - conditioned STI symptoms | 290 | -0.036** | -0.072 - -0.000 | 290 | -0.036* | -0.072 - 0.001 |
| HIV - removed STI symptoms | 284 | -0.034** | -0.065 - -0.003 | 284 | -0.035** | -0.066 - -0.005 |

Notes: Unadjusted = Ordinary Least Squares (OLS) regression of follow-up data. Adjusted = OLS regression adjusting treatment effect for baseline levels of outcomes, if the outcome was collected during midline, then baseline controls, namely log of adult equivalent expenditure, number of economic dependents, household size and if the respondent has a child. Maximum likelihood estimation used for logistic regression did not converge therefore these models are estimated with OLS which gives less robust estimates in this situation and why we illustrate the findings descriptively

**Table A4:** Difference between baseline characteristics of attritors in the commercial sex strata.

| Characteristics | Mean of control attritors | Mean of treatment attritors | Difference | p-val |
| --- | --- | --- | --- | --- |
| Age, years | 26.778 | 28.683 | 1.905 | 0.270 |
| Experience - months in sex work | 38.021 | 49.214 | 11.193 | 0.133 |
| **Nationality** |  |  |  |  |
| Cameroonian (%) | 1.000 | 1.000 | 0.000 | - |
| **Education** |  |  |  |  |
| Currently studying | 0.191 | 0.214 | 0.023 | 0.792 |
| **Highest achieved level of education** |  |  |  |  |
| Primary | 0.170 | 0.143 | -0.027 | 0.727 |
| Secondary primary cycle | 0.319 | 0.333 | 0.014 | 0.888 |
| Secondary second cycle | 0.255 | 0.238 | -0.017 | 0.853 |
| Superior | 0.213 | 0.238 | 0.025 | 0.778 |
| No education | 0.043 | 0.048 | 0.005 | 0.910 |
| Other | 0.000 | 0.000 | 0.000 | - |
| **Marital status** |  |  |  |  |
| Never married | 0.936 | 0.881 | -0.055 | 0.369 |
| Separated | 0.021 | 0.071 | 0.050 | 0.259 |
| Divorced | 0.043 | 0.024 | -0.019 | 0.629 |
| Widowed | 0.000 | 0.024 | 0.024 | 0.293 |
| **Economic Variables** |  |  |  |  |
| Household size | 2.617 | 2.762 | 0.145 | 0.680 |
| Number of economic dependents | 0.851 | 0.786 | -0.065 | 0.429 |
| Number that have child economic dependent | 1.617 | 1.929 | 0.312 | 0.363 |
| Number of children | 3.745 | 3.976 | 0.232 | 0.649 |
| Household expenditure per adult equivalent | 53,625.969 | 71,589.680 | 17,963.713 | 0.473 |
| Observations | 47 | 42 | 89 (total) |  |

**Table A5:** Difference between baseline characteristics of attritors in the transactional sex strata.

| Characteristics | Mean of control attritors | Mean of treatment attritors | Difference | p-val |
| --- | --- | --- | --- | --- |
| Age, years | 22.182 | 24.148 | 1.966 | 0.288 |
| Experience - months in sex work | 46.909 | 32.577 | -14.332 | 0.276 |
| **Nationality** |  |  |  |  |
| Cameroonian (%) | 0.909 | 1.000 | 0.091 | 0.118 |
| **Education** |  |  |  |  |
| Currently studying | 0.182 | 0.444 | 0.263 | 0.135 |
| **Highest achieved level of education** |  |  |  |  |
| Primary | 0.000 | 0.074 | 0.074 | 0.367 |
| Secondary primary cycle | 0.545 | 0.333 | -0.212 | 0.236 |
| Secondary second cycle | 0.364 | 0.296 | -0.067 | 0.695 |
| Superior | 0.091 | 0.222 | 0.131 | 0.357 |
| No education | 0.000 | 0.000 | 0.000 | - |
| Other | 0.000 | 0.074 | 0.074 | 0.367 |
| **Marital status** |  |  |  |  |
| Never married | 1.000 | 1.000 | 0.000 | - |
| Separated | 0.000 | 0.000 | 0.000 | - |
| Divorced | 0.000 | 0.000 | 0.000 | - |
| Widowed | 0.000 | 0.000 | 0.000 | - |
| **Economic Variables** |  |  |  |  |
| Household size | 3.364 | 2.926 | -0.438 | 0.429 |
| Number of economic dependents | 0.818 | 0.889 | 0.071 | 0.571 |
| Number that have child economic dependent | 1.636 | 1.222 | -0.414 | 0.451 |
| Number of children | 5.273 | 4.630 | -0.643 | 0.437 |
| Household expenditure per adult equivalent | 29,996.332 | 30,686.875 | 690.544 | 0.916 |
| Observations | 11 | 27 | 38 (total) |  |

**Table A6:** Odds ratios for health utilisation outcomes.

| Outcomes | n | Unadjusted odds |  | n | Adjusted odds |  |
| --- | --- | --- | --- | --- | --- | --- |
|  |  | OR | 95% CI |  | AOR | 95% CI |
| **Commercial Sex** |  |  |  |  |  |  |
| **Binary outcome** |  |  |  |  |  |  |
| If sick since randomisation | 489 | 1.314 | 0.920 - 1.877 | 489 | 1.317 | 0.920 - 1.886 |
| If respondent sought a health professional | 228 | 1.287 | 0.662 - 2.502 | 228 | 1.230 | 0.624 - 2.426 |
| If at least 1 economic dependent reported sick | 489 | 0.789 | 0.553 - 1.126 | 489 | 0.785 | 0.540 - 1.142 |
| Suffered health expense shock in the last 30 days | 489 | 0.671** | 0.457 - 0.986 | 489 | 0.651** | 0.441 - 0.961 |
| **Transactional Sex** |  |  |  |  |  |  |
| **Binary outcome** |  |  |  |  |  |  |
| If sick since randomisation | 530 | 1.133 | 0.805 - 1.595 | 530 | 1.143 | 0.809 - 1.616 |
| If respondent sought a health professional | 290 | 2.131*** | 1.198 - 3.790 | 290 | 2.327*** | 1.279 - 4.233 |
| If at least 1 economic dependent reported sick | 530 | 0.873 | 0.617 - 1.237 | 530 | 0.975 | 0.676 - 1.405 |
| Suffered health expense shock in the last 30 days | 530 | 0.765 | 0.521 - 1.125 | 530 | 0.790 | 0.533 - 1.169 |
| **Pooled (commercial + transactional)** |  |  |  |  |  |  |
| **Binary outcome** |  |  |  |  |  |  |
| If sick since randomisation | 1,019 | 1.212 | 0.948 - 1.550 | 1,019 | 1.220 | 0.953 - 1.563 |
| If respondent sought a health professional | 518 | 1.729** | 1.122 - 2.664 | 518 | 1.741** | 1.121 - 2.703 |
| If at least 1 economic dependent reported sick | 1,019 | 0.831 | 0.649 - 1.063 | 1,019 | 0.872 | 0.674 - 1.128 |
| Suffered health expense shock in the last 30 days | 1,019 | 0.718** | 0.547 - 0.942 | 1,019 | 0.727** | 0.553 - 0.957 |

Notes: Unadjusted = logistic regression of follow-up data. Adjusted = Logistic regression adjusting treatment effect for baseline levels of outcomes, if the outcome was collected during midline, then baseline controls, namely log of adult equivalent expenditure, number of economic dependents, household size, if the respondent has a child and if the respondent is enrolled in school. The recall period for if the respondent or dependent was sick and if they saw a health professional is "last 30 days" in the baseline but "since randomisation" in the midline and endline.

**Table A7:** Odds ratio of behavioural outcomes.

| Sexual Behaviour Outcomes | Unadjusted odds |  |  | Adjusted odds |  |  |
| --- | --- | --- | --- | --- | --- | --- |
|  | n | OR | 95% CI | n | AOR | 95% CI |
| **Commercial Sex** |  |  |  |  |  |  |
| Used condom at last sex act - Direct questioning | 364 | 1.151 | 0.600 - 2.207 | 364 | 1.148 | 0.589 - 2.238 |
| Quit commercial sex work | 439 | 1.045 | 0.666 - 1.640 | 439 | 1.055 | 0.655 - 1.701 |
| Quit commercial sex work - bootstrapped | 439 | 1.045 | 0.672 - 1.625 | 439 | 1.055 | 0.651 - 1.712 |
| **Transactional Sex** |  |  |  |  |  |  |
| Used condom at last sex act - Direct questioning | 399 | 1.099 | 0.738 - 1.635 | 399 | 1.069 | 0.712 - 1.604 |
| Quit transactional sex | 496 | 1.687** | 1.089 - 2.615 | 496 | 1.682** | 1.075 - 2.632 |
| Quit transactional sex - bootstrapped | 496 | 1.687** | 1.090 - 2.614 | 496 | 1.682** | 1.042 - 2.715 |
| **Pooled (commercial + transactional)** |  |  |  |  |  |  |
| Used condom at last sex act - Direct questioning | 763 | 1.149 | 0.853 - 1.548 | 763 | 1.189 | 0.869 - 1.627 |
| Number exited either type of sex work | 935 | 1.340* | 0.980 - 1.832 | 935 | 1.308 | 0.948 - 1.804 |
| Number exited either type of sex work - bootstrapped | 935 | 1.340* | 0.963 - 1.863 | 935 | 1.308* | 0.957 - 1.787 |

Notes: Unadjusted = logistic regression of follow-up data. Adjusted = Logistic regression adjusting treatment effect for baseline levels of outcomes, if the outcome was collected during midline, then baseline controls, namely log of adult equivalent expenditure, number of economic dependents, household size, if the respondent has a child and if the respondent is enrolled in school. The list experiment was estimated using OLS using the double list experiment method (Lépine et al., 2020) hence twice the observations as each respondent appears twice.

**Table A8:**
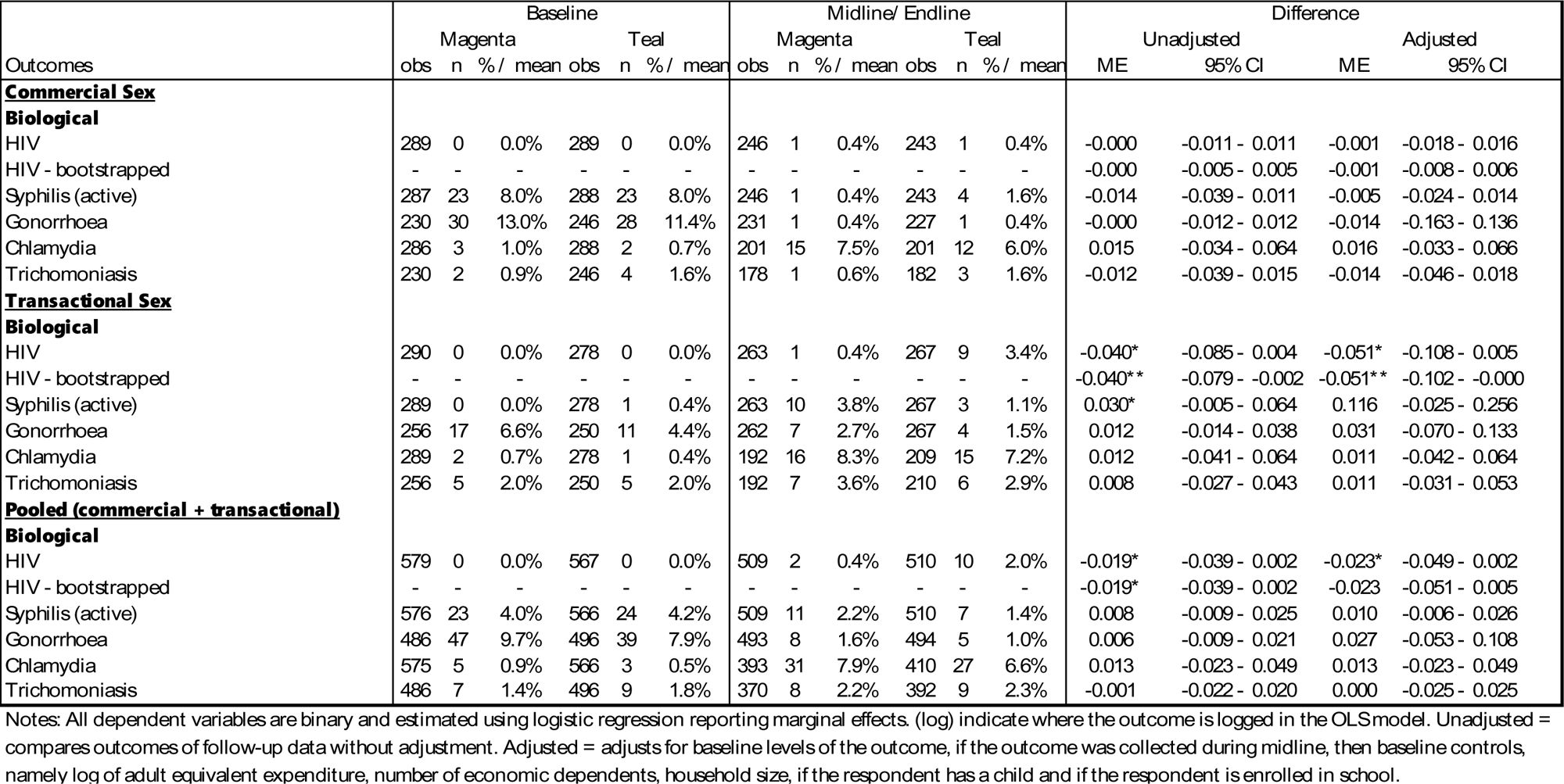
Biological outcomes reporting their marginal effects.

| Outcomes | Baseline |  |  |  |  |  | Midline/ Endline |  |  |  |  |  | Difference |  |  |  |
| --- | --- | --- | --- | --- | --- | --- | --- | --- | --- | --- | --- | --- | --- | --- | --- | --- |
|  | Magenta |  |  | Teal |  |  | Magenta |  |  | Teal |  |  | Unadjusted |  | Adjusted |  |
|  | obs | n | % / mean | obs | n | % / mean | obs | n | % / mean | obs | n | % / mean | ME | 95% CI | ME | 95% CI |
| **Commercial Sex** |  |  |  |  |  |  |  |  |  |  |  |  |  |  |  |  |
| **Biological** |  |  |  |  |  |  |  |  |  |  |  |  |  |  |  |  |
| HIV | 289 | 0 | 0.0% | 289 | 0 | 0.0% | 246 | 1 | 0.4% | 243 | 1 | 0.4% | -0.000 | -0.011 - 0.011 | -0.001 | -0.018 - 0.016 |
| HIV - bootstrapped | - | - | - | - | - | - | - | - | - | - | - | - | -0.000 | -0.005 - 0.005 | -0.001 | -0.008 - 0.006 |
| Syphilis (active) | 287 | 23 | 8.0% | 288 | 23 | 8.0% | 246 | 1 | 0.4% | 243 | 4 | 1.6% | -0.014 | -0.039 - 0.011 | -0.005 | -0.024 - 0.014 |
| Gonorrhoea | 230 | 30 | 13.0% | 246 | 28 | 11.4% | 231 | 1 | 0.4% | 227 | 1 | 0.4% | -0.000 | -0.012 - 0.012 | -0.014 | -0.163 - 0.136 |
| Chlamydia | 286 | 3 | 1.0% | 288 | 2 | 0.7% | 201 | 15 | 7.5% | 201 | 12 | 6.0% | 0.015 | -0.034 - 0.064 | 0.016 | -0.033 - 0.066 |
| Trichomoniasis | 230 | 2 | 0.9% | 246 | 4 | 1.6% | 178 | 1 | 0.6% | 182 | 3 | 1.6% | -0.012 | -0.039 - 0.015 | -0.014 | -0.046 - 0.018 |
| **Transactional Sex** |  |  |  |  |  |  |  |  |  |  |  |  |  |  |  |  |
| **Biological** |  |  |  |  |  |  |  |  |  |  |  |  |  |  |  |  |
| HIV | 290 | 0 | 0.0% | 278 | 0 | 0.0% | 263 | 1 | 0.4% | 267 | 9 | 3.4% | -0.040* | -0.085 - 0.004 | -0.051* | -0.108 - 0.005 |
| HIV - bootstrapped | - | - | - | - | - | - | - | - | - | - | - | - | -0.040** | -0.079 - -0.002 | -0.051** | -0.102 - -0.000 |
| Syphilis (active) | 289 | 0 | 0.0% | 278 | 1 | 0.4% | 263 | 10 | 3.8% | 267 | 3 | 1.1% | 0.030* | -0.005 - 0.064 | 0.116 | -0.025 - 0.256 |
| Gonorrhoea | 256 | 17 | 6.6% | 250 | 11 | 4.4% | 262 | 7 | 2.7% | 267 | 4 | 1.5% | 0.012 | -0.014 - 0.038 | 0.031 | -0.070 - 0.133 |
| Chlamydia | 289 | 2 | 0.7% | 278 | 1 | 0.4% | 192 | 16 | 8.3% | 209 | 15 | 7.2% | 0.012 | -0.041 - 0.064 | 0.011 | -0.042 - 0.064 |
| Trichomoniasis | 256 | 5 | 2.0% | 250 | 5 | 2.0% | 192 | 7 | 3.6% | 210 | 6 | 2.9% | 0.008 | -0.027 - 0.043 | 0.011 | -0.031 - 0.053 |
| **Pooled (commercial + transactional)** |  |  |  |  |  |  |  |  |  |  |  |  |  |  |  |  |
| **Biological** |  |  |  |  |  |  |  |  |  |  |  |  |  |  |  |  |
| HIV | 579 | 0 | 0.0% | 567 | 0 | 0.0% | 509 | 2 | 0.4% | 510 | 10 | 2.0% | -0.019* | -0.039 - 0.002 | -0.023* | -0.049 - 0.002 |
| HIV - bootstrapped | - | - | - | - | - | - | - | - | - | - | - | - | -0.019* | -0.039 - 0.002 | -0.023 | -0.051 - 0.005 |
| Syphilis (active) | 576 | 23 | 4.0% | 566 | 24 | 4.2% | 509 | 11 | 2.2% | 510 | 7 | 1.4% | 0.008 | -0.009 - 0.025 | 0.010 | -0.006 - 0.026 |
| Gonorrhoea | 486 | 47 | 9.7% | 496 | 39 | 7.9% | 493 | 8 | 1.6% | 494 | 5 | 1.0% | 0.006 | -0.009 - 0.021 | 0.027 | -0.053 - 0.108 |
| Chlamydia | 575 | 5 | 0.9% | 566 | 3 | 0.5% | 393 | 31 | 7.9% | 410 | 27 | 6.6% | 0.013 | -0.023 - 0.049 | 0.013 | -0.023 - 0.049 |
| Trichomoniasis | 486 | 7 | 1.4% | 496 | 9 | 1.8% | 370 | 8 | 2.2% | 392 | 9 | 2.3% | -0.001 | -0.022 - 0.020 | 0.000 | -0.025 - 0.025 |

Notes: All dependent variables are binary and estimated using logistic regression reporting marginal effects. (log) indicate where the outcome is logged in the OLS model. Unadjusted = compares outcomes of follow-up data without adjustment. Adjusted = adjusts for baseline levels of the outcome, if the outcome was collected during midline, then baseline controls, namely log of adult equivalent expenditure, number of economic dependents, household size, if the respondent has a child and if the respondent is enrolled in school.

#### 7.2 List experiment method and double list experiment

Our primary measure of risky sexual behaviour within sex acts is condom use collected via the verified double list experiment method, an indirect elicitation method. The advantage of the list experiment is that it allows respondents in our survey to answer sensitive questions, in our case about the use of condoms during their last sex act, allowing confidentiality since the enumerator or researchers are not able to assign the behaviour to a specific individual. It allows a more accurate prevalence of condomless sex to be estimated, minimising social desirability bias. Previously, the list experiment method has been used for eliciting abortion (Moseson et al., 2021; Bell and Bishai, 2019), voting preferences (Gonzalez-Ocantos et al., 2012; Holbrook and Krosnick, 2010), use of micro-finance loans (Karlan and Zinman, 2012), opinions on undocumented migrants (McKenzie and Siegel, 2013), gay marriage (Lax et al., 2016) and racism (Krumpal, 2013) and has been proven to be effective to measure condom use (LaBrie and Earleywine, 2000; Treibich and Lépine, 2019). There is debate over the effectiveness of the list experiment in measuring sensitive behaviours. Lensvelt-Mulders et al. (2016) performed a meta-analysis finding it more accurate than face-to-face questioning at estimating prevalence of sensitive behaviours, whereas several other studies find issues often resulting from a poor list experiment design, including respondents’ understanding is high (Haber et al., 2018) and Imai (2012) and Imai (2011).

##### 7.2.1 Implementation

Here we describe the statements for the women engaging in transactional sex, but the method is identical for the commercial sex group. During the survey, when an enumerator reaches the list experiment question, their respondent is randomly allocated to the treatment or control groups for the list experiment and asked how many of the following statements the respondent agrees with. It then lists either three non-sensitive statements for the control group:

- Usually, I meet my sugar daddies on the street.
- My sugar daddy is older than me.
- Monday is the day I see most of my sugar daddies.

Or for the treatment group, it lists the same three non-sensitive statements plus a sensitive statement of interest in position 2:

- Usually, I meet my sugar daddies on the street.
- **I used a condom the last time I had sex with my last sugar daddy.**
- My sugar daddy is older than me.
- Monday is the day I see most of my sugar daddies.

The key assumption is that the average number of non-sensitive statements agreed with is the same for the treatment and control groups. Therefore the difference in the average number of statements agreed with is the prevalence of condom use at the last sex act.

The double list experiment method simply repeats the list experiment with a new set of non-sensitive statements and reverses the treatment and control groups allocated in the first experiment. This means each respondent receives the sensitive statement at least once during the interview. The second set of non-sensitive statements are:

- I like to look good before I go out.
- I consider myself as a sex worker.
- I want to marry my sugar daddy.

The prevalence can also be estimated using OLS regression analysis. When using the double list experiment, each respondent appears in the model as two observations, one when they were in the control group and one in the treatment group of the list experiment.

As you can see, the advantage of this method is that there is no way for the researcher to back out the true answer to the sensitive statement that a respondent has, providing privacy to answer in confidence. This strength is also a drawback since interpretation of findings can only be made about a groups prevalence and not at the individual level. The validity of the list experiment has been examined and proven elsewhere (31,36).

For the commercial sex arm the two sets of non-sensitive statements are:

- Usually, I meet my clients at the couloir.
- I prefer the client to pay me before the act.
- Monday is the day I have the most clients.

And,

- I like to look good before I go out.
- I like all of my clients.
- Usually, I solicit clients by phone.

The sensitive statement is worded as:

- I used a condom the last time I had sex with my last client.

##### 7.2.2 Estimating equation

The prevalence can almost be estimated using simple OLS regression that can also be used to estimate the difference in the prevalence between sub-samples within our dataset. To determine the impact of the intervention on condom use collected via the list experiment, we estimate the following equation:

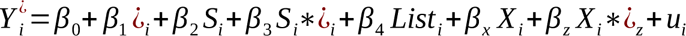

Where *Y* indicates the number of statements the individual *i,* agreed with during the list experiment (*LE*). ¿ indicates if the individual received the treatment or control list, *S* indicates if the respondent was part of the treatment group or not. Since we are using the validated double list experiment where each individual is asked the list experiment twice, once as a treatment list and once as a control list, the term *List* is an indicator for which set of non-sensitive statements they received. *X* is a list of individual controls. Models are estimated with robust standard errors clustered at the individual since each respondent is included twice within each model, once for the first instance of the list experiment and once for the second instance. *β*_3_ is our parameter of interest and represents the difference in condom use between the treated and control groups or the marginal impact of the intervention on condom use at the last sex act.

#### 7.3 STI measurement issues

Below are further details about the lack of effects found for STIs other than HIV.

For gonorrhoea and trichomoniasis, tests required vaginal swabs that are difficult to collect (37) and testing in the lab, meaning there was a high rate of refusal (gonorrhoea and trichomoniasis refusal rate was 17% at baseline, 14% at midline and 10% at the endline vs. zero refusals for HIV across all surveys). We discovered there were issues with transportation and storage in hot and humid environments that impacted the accuracy of the tests because samples needed to be cold stored (24). We found zero cases of gonorrhoea at the endline^4^, yet an average prevalence of 9.2% at midline. Trichomoniasis results did not show different results between midline and endline, but since the samples tested were the same as gonorrhoea, they suffered the same refusal and transportation issues, limiting their validity.

During baseline and midline surveys, every 15th chlamydia, syphilis and HIV test performed (including HIV), plus all inconclusive and positive tests were sent to a lab for verification using the same rapid test as was performed on-site. We found many of the positive tests taken during the survey came back negative from the lab. Therefore at endline, re-testing was moved to the ELISA laboratory where a new chlamydia test and more robust syphilis test were performed. We then found that many of the 15th sample re-tests were coming back positive after a negative rapid test. Upon this realisation, we sent all chlamydia and syphilis samples for lab testing as well as completing the rapid tests at the survey site. This led to our findings of poor sensitivity and specificity of chlamydia and syphilis tests currently recommended by the Cameroonian Government.

#### 7.4 Healthcare utilisation and out-of-pocket expenses

Previous literature has not always found insurance reduces OOP significantly for a variety of reasons, including informal payments and drug stockouts (38,39). The intervention was designed in order to minimise the chance of or level of OOP payments that could be made by individuals accessing services. On average, 70% of insured participants or their economic dependents were sick at least once over the treatment period. 57% of participants reported to have received free care as a result of the intervention for themselves or their economic dependents. There were significant reductions in the chance of suffering a health expense shock and significant and large reductions in the level of medical out-of-pocket spending for the treatment group. Among those who incurred some expenses, the average OOPs was only 877 CFA (£1.12) per visit^5^ (in comparison to 30,250 CFA (£38.72) incurred by the control group during their last illness^6^).

To ensure health care quality was not compromised, weekly reviews of medical cases with a senior medical doctor independent from the hospital used for the intervention were done. These measures ensured that everyone attending the intervention received the best care possible and that the hospital was not cutting corners. There were no issues reported as a result of these reviews.

In addition, we conducted a patient survey and in-depth interviews with patients to measure their overall satisfaction. Results indicated that patients were satisfied by the care received and that there was no informal payments. The only issue reported was the drug stockouts for some essential medicines.

##### 7.4.1 Expense shocks or health treatment

The impact of the intervention on HIV for women in transactional sex can work through two channels, see Figure 1. First, through protection against economic shocks that reduce the incentives to engage in risky behaviours resulting in less risky sex; second, through treatment for STIs and therefore lower susceptibility to HIV that way (29). To somewhat disentangle these two mechanisms, we can examine the role of self-reported STI symptoms in mediating the intervention’s impact on HIV in the transactional arm.

Another method to check the channel the intervention works is to examine those who are expected to benefit more or less from the intervention. Whilst predicting illness is impossible, we know that those with more economic dependents will have a greater chance of both suffering an illness and receiving free care for the illness. Therefore, we test whether the intervention impacted condom use (using the list experiment) for those with more or less than three economic dependents finding the intervention leads to a 25% increase in condom use (ME=0.248, p<0.05) for women in transactional sex with more than three economic dependents but has no effect (ME=-0.067, p>0.1) for those with less than three economic dependents^7^.

Together this evidence suggests the primary mechanism at work is that of protection against economic shocks rather than through treatment of STIs.

#### 7.5 Moral Hazard

The intervention succeeded in its primary aims of increasing the likelihood of reducing the costs of seeking health care when respondents or economic dependents were sick or injured. The likelihood of suffering a health-related economic shock also fell alongside typical and last 30-day health-related out-of-pocket payments, see Table 5. Moral hazard could counter the reductions in risk through protection from the intervention. Ex-ante recipients could increase risky health behaviours because of health insurance provision, or ex-post are more likely to use more healthcare than needed after an accident or illness occurs. The latter is difficult to measure and is not relevant to our mechanism. However, increased risk taking behaviour in the knowledge treatment is provided for free could counteract reduction in risks from shocks protection.

Table 5 shows no difference in reported illness between treatment and control groups at follow-up, suggesting little moral hazard in reporting of illness or increased risk-taking leading to illness overall. We also test moral hazard by comparing risky behaviours of those who do not report an illness or injury to themselves or an economic dependent; since they did not need the financial protection, any differences are likely to suggest if moral hazard is occurring. Table A1 shows no increase in risky behaviours due to the treatment for those without the need to use it. In fact, treatment reduces risky behaviours in these people with protected sex increasing (ME=0.196, p<0.05, transactional only), and oral sex acts falling (ME=-0.085, p<0.05, pooled) among women who did not report any illness.

## Footnotes

1 As of 18^th^ August 2022

2 We include self-reported UTIs as well since these have similar symptoms to some STIs and might have been mischaracterised be respondents.

3 There were significantly fewer STI symptoms reported in the treatment groups giving further evidence of the intervention protecting against risky behaviours.

4 Those positive cases in Table A0 & Table A8 were all recorded at midline and differential refusal rates mean there are more test results in our analysis sample than the baseline sample.

5 Collected via cost paid by hospital logs.

6 Collected via the survey. The comparative value for the treatment group was 15150 CFAF, around half of the control group, but includes non-covered illnesses that would not have been entered into the hospital log.

7 Not reported in tables.

## Notes

### Competing Interest Statement

The authors have declared no competing interest.

### Clinical Trial

ISRCTN22516548

### Funding Statement

The study is funded by the UKRI Future Leaders Fellowship awarded to Dr Lepine

### Author Declarations

UCL ethics committee and national ethics committee in Cameroon

